# Impact of COVID-19 vaccination on symptoms and immune phenotypes in vaccine-naïve individuals with Long COVID

**DOI:** 10.1101/2024.01.11.24300929

**Authors:** Connor B Grady, Bornali Bhattacharjee, Julio Silva, Jillian Jaycox, Lik Wee Lee, Valter Silva Monteiro, Mitsuaki Sawano, Daisy Massey, César Caraballo, Jeff R. Gehlhausen, Alexandra Tabachnikova, Tianyang Mao, Carolina Lucas, Mario A. Peña-Hernandez, Lan Xu, Tiffany J. Tzeng, Takehiro Takahashi, Jeph Herrin, Diana Berrent Güthe, Athena Akrami, Gina Assaf, Hannah Davis, Karen Harris, Lisa McCorkell, Wade L Schulz, Daniel Grffin, Hannah Wei, Aaron M Ring, Leying Guan, Charles Dela Cruz, Akiko Iwasaki, Harlan M Krumholz

## Abstract

**Background:** Long COVID contributes to the global burden of disease. Proposed root cause hypotheses include the persistence of SARS-CoV-2 viral reservoir, autoimmunity, and reactivation of latent herpesviruses. Patients have reported various changes in Long COVID symptoms after COVID-19 vaccinations, leaving uncertainty about whether vaccine-induced immune responses may alleviate or worsen disease pathology.

**Methods:** In this prospective study, we evaluated changes in symptoms and immune responses after COVID-19 vaccination in 16 vaccine-naïve individuals with Long COVID. Surveys were administered before vaccination and then at 2, 6, and 12 weeks after receiving the first vaccine dose of the primary series. Simultaneously, SARS-CoV-2-reactive TCR enrichment, SARS-CoV-2-specific antibody responses, antibody responses to other viral and self-antigens, and circulating cytokines were quantified before vaccination and at 6 and 12 weeks after vaccination.

**Results:** Self-report at 12 weeks post-vaccination indicated 10 out of 16 participants had improved health, 3 had no change, 1 had worse health, and 2 reported marginal changes. Significant elevation in SARS-CoV-2-specific TCRs and Spike protein-specific IgG were observed 6 and 12 weeks after vaccination. No changes in reactivities were observed against herpes viruses and self-antigens. Within this dataset, higher baseline sIL-6R was associated with symptom improvement, and the two top features associated with non-improvement were high IFN-β and CNTF, among soluble analytes.

**Conclusions:** Our study showed that in this small sample, vaccination improved the health or resulted in no change to the health of most participants, though few experienced worsening. Vaccination was associated with increased SARS-CoV-2 Spike protein-specific IgG and T cell expansion in most individuals with Long COVID. Symptom improvement was observed in those with baseline elevated sIL-6R, while elevated interferon and neuropeptide levels were associated with a lack of improvement.

**Plain language summary:** The impact of the COVID-19 vaccine on vaccine-naïve individuals suffering from Long COVID is uncertain. This study assessed the experience and immune signatures of 16 unvaccinated participants with Long COVID. A total of 10 participants had improved health status after vaccination, and one person reported only worsening health. As expected, vaccination increased immune cells and antibodies against the viral spike protein. Immune signatures may prove to be predictors of health status after vaccination. However, given the small number of participants, these initial findings need further validation.

## Introduction

Long COVID, also known as post-acute sequelae of SARS-CoV-2 infection (PASC), is a debilitating condition following acute SARS-CoV-2 infection.^1–6^ It can significantly impact people’s lives, including their ability to return to work and engage in other social activities.^7,8^ Although investigators have launched several prospective clinical trials of Long COVID treatment,^9–12^ no definitive therapies exist.

Viral persistence is a possible contributing factor for Long COVID.^13–15^ So far, several reports have suggested the persistence of active SARS-CoV-2 reservoirs in Long COVID patients^16,17^ and that vaccination could assist in clearing persistent virus. However, the impact of vaccination after developing Long COVID remains unclear.^18^ In a recent study, Nayyerabadi et al. reported the alleviation of symptoms, an increase in WHO-5 well-being scores, and a decrease in inflammatory cytokines after vaccination among participants with Long COVID.^19^

At the same time, there are concerns that the vaccine’s spike protein or innate immune stimuli induced by the lipid nanoparticles and mRNA may exacerbate Long COVID symptoms by activating immunological pathways.^14,20^ These concerns have contributed to vaccine hesitancy among individuals with Long COVID.^21^ Consequently, it is crucial to investigate the effect of vaccination on Long COVID symptoms.

Accordingly, we launched the Yale COVID-19 Recovery Vaccine Study: Measuring Changes in Long Covid Symptoms After Vaccination (NCT04895189), a prospective, unblinded, observational study to evaluate changes in Long COVID symptoms, their prevalence, and burden. Immune responses before and after receiving the COVID-19 vaccine were evaluated to assess vaccine responses and identify factors associated with health outcomes in vaccine-naïve individuals with Long COVID.^22^ This report presents findings from 16 participants recruited between May 3, 2021, and February 2, 2022.

## Methods

### Study design

A pre-post, prospective observational study was conducted among unvaccinated individuals experiencing Long COVID symptoms who intended to receive a COVID-19 vaccine as part of the routine clinical care (ClinicalTrials.gov Identifier: NCT04895189). Participants completed a survey before vaccination to collect demographic and acute COVID-19 infection information and their baseline (i.e., pre-vaccination) Long COVID symptom experience (survey included in the **Supplementary file**). Participants were vaccinated with any approved COVID-19 vaccine and then asked to complete three follow-up surveys at 2, 6, and 12 weeks after receiving the first vaccine dose of the primary series. SARS-CoV-2 specific humoral responses, responses to common viral pathogens and autoantigens, T-cell repertoire sequencing, and soluble immune modulators were quantified in a subset of participants before vaccination and at 6 (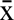 : 6.6) and 12 (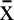 : 13.4) weeks after vaccination. This study was approved by the Yale University Institutional Review Board (IRB #2000030423).

### Patient involvement

Patient advocacy groups were actively engaged in the conception and design of the study. The idea for the study originated from a Survivor Corps poll posted to their Facebook page, many of whom had COVID-19 and suffered from Long COVID. Their poll showed that 40% of respondents with self-reported Long COVID had mild to full symptom resolution after vaccination while 14% reported worsening of their symptoms. In response, hypotheses were developed as to how vaccination might impact Long COVID symptoms.^23,24^ Survivor Corps aided in participant recruitment. The Patient-Led Research Collaborative, a self-organized group of Long COVID patient-researchers working on patient-led research around the Long COVID experience, was enlisted to contribute to the study design. Both groups advocated for including individuals without a positive PCR test for SARS-CoV-2 and helped develop study surveys. Surveys were also informed by prior survey studies.^7,25–29^

### Eligibility

Eligible individuals included unvaccinated individuals 12 years or older who self-reported Long COVID (based on the presence of symptoms that started after COVID-19 and persisted more than two months) and planned to receive the COVID-19 vaccine. To verify past COVID-19 illness, individuals must have had a positive COVID-19 test (PCR or antigen) more than two months prior, have had a positive COVID-19 antibody or T-cell test, have been hospitalized for COVID-19, or have been diagnosed by a clinician as having COVID-19. Participants also had to be willing to travel to New Haven, Connecticut to provide blood and saliva samples. Recruitment was conducted through social media advertisements and patient support groups. Participants were not compensated for their involvement.

### Outcomes

The primary outcome was whether individuals’ overall health condition improved, stayed the same, or worsened after receiving a COVID-19 vaccine. Secondary outcomes included changes in symptom prevalence and severity and associated changes in immune response to the COVID-19 vaccine. The immunophenotyping assays included the detection of SARS-CoV-2-specific antibody responses, SARS-CoV-2 specific T cell enrichment, antibody responses to other common viruses and quantitation of soluble immune mediators.

### Data collection

Before vaccination, demographic, acute COVID-19, and persistent symptom information was collected by survey. Participants were asked to rate their symptoms in terms of how much physical pain or discomfort the symptom caused (“physical effects”) and how much each symptom impaired their social or family functioning compared to before infection (“social effects”) on a 5-level Likert scale from “not at all” to “very much” (**Supplementary table 1**). We provided a list of 125 symptoms developed through a literature review and prior Long COVID symptoms lists.^7,8,26^ Participants were asked the same questions on the three post-vaccination surveys (2, 6, and 12 weeks after vaccination). Overall health change was measured with the question, “Would you say that your overall health, as compared to your health before the vaccine, is worse, better, or the same?” at 2, 6, and 12 weeks after vaccination. Data collection was performed using RedCap version 12.0.25 (Vanderbilt University). All survey data were self-reported. Blood samples were collected on-site before vaccination and at 6 and 12 weeks after vaccination. Further information on the study’s design, eligibility criteria, and data collection are available online.^22^

### Biospecimen processing

Whole blood was collected in sodium-heparin-coated vacutainers (BD 367874, BD Biosciences) and EDTA-coated vacutainers (BD 367856, BD Biosciences). For each participant, unique study identifiers were provided upon collection. Plasma samples were collected after centrifugation of whole blood at 600×g for 10 minutes at room temperature (RT) without brake from sodium-heparin-coated tubes as previously described.^30^ The blood samples collected in EDTA-coated tubes were frozen and subsequently shipped to Adaptive Biotechnologies for TCR sequencing.

### Quantitation of SARS-CoV-2 specific antibody levels by ELISA

ELISA assays were performed as previously described.^30^ Briefly, MaxiSorp plates (96 wells; 442404, Thermo Scientific) were coated with recombinant SARS-CoV-2 S1 (S1N-C52H3-100 μg, ACROBiosystems), receptor-binding domain (RBD) (SPD-C52H3-100 μg, ACROBiosystems) and the nucleocapsid protein (NUN-C5227-100 μg, ACROBiosystems) at a concentration of 2 μg/ml in PBS and were incubated overnight at 4 °C. The primary antibodies used for the standard curves were human anti-spike (SARS-CoV-2 human anti-spike [AM006415; 91351, Active Motif]) and human anti-nucleocapsid (SARS-CoV-2 anti-nucleocapsid [1A6; MA5-35941, Active Motif]) and HRP anti-human IgG antibody (1:5,000; A00166, GenScript) was the secondary antibody.

### TCR sequencing & SARS-CoV-2 Specific TCR assignment

Immunosequencing of the third complementarity determining (CDR3) regions of TCR-β chains was carried out using ImmunoSEQ Assays (Adaptive Biotechnologies). Samples were classified as positive or negative for detection and enrichment of COVID-specific T cells using four of Adaptive’s COVID-19 classifiers: V1 classifier, V3 classifier, spike classifier and non-spike classifier. The V1 classifier was trained comparing peripheral repertoires from acute COVID and convalescent subjects with control samples collected pre-pandemic.^31,32^ The V3 classifier was trained on a larger dataset that included subjects with natural infection as well as those that were vaccinated as positive cases. The sequences in the V3 classifier were cross-referenced against data from MIRA (multiplexed antigen-stimulation experiments) experiment to develop two additional classifier.^32,33^ The spike classifier identifies the spike-specific signal while the non-spike classifier (with vaccinated samples included as controls) identifies natural infection using the non-spike signal. T cell responses are categorized as negative, positive, and “No Call” (representing samples with an insufficient number of T cell rearrangements to make a definitive negative call).

### Rapid Extracellular Antigen Profiling (REAP) Library Expansion

The new yeast library (Exo205) containing 6,452 unique antigens was used. IgG isolations and REAP selections were done as previously described.^30^ Briefly, participant IgGs were purified from plasma using protein G magnetic beads and yeast-reactive IgGs were initially removed by adsorption to yeast transformed with the pDD003 empty vector. A total of 10^8^ induced Exo205 yeast cells were washed with PBE and incubated with 10 μg of purified participant IgGs in duplicate. IgG bound yeast cells were selected by anti-human IgG Fc antibody binding (clone QA19A42, Biolegend) and next generation sequencing (NGS) was carried out to identify epitopes based on the protein display barcode on yeast plasmids. REAP scores were calculated as described previously.^30^

### Multiplex proteomic analysis

Frozen patient plasma was shipped to Eve Technologies (Calgary, Alberta, Canada) on dry ice to run 13 multiplex panels: Human Cytokine/Chemokine 71-plex Discovery Assay (HD71), Human Cytokine P3 Assay (HCYP3-07), Human Cytokine Panel 4 Assay (HCYP4-19), Human Complement Panel Assay (HDCMP1), Human Myokine Assay (HMYOMAG-10), Human Neuropeptide Assay (HNPMAG-05), Human Pituitary Assay (HPTP1), Human Adipokine Panel 2 Assay (HADK2-03), Human Cardiovascular Disease Panel Assay (HDCVD9), Human CVD2 Assay (HCVD2-8), Steroid/Thyroid 6plex Discovery Assay (STTHD) Human Adipokine Assay (HDADK5), and TGF-Beta 3-plex Discovery Assay (TGFβ1-3). Samples were sent in two batches with internal controls in each shipment to assess effectiveness of batch correction as described below.

To harmonize data across the two batches, ComBat was used, an empirical Bayes method available through the “sva”^34^ R package (version 3.4.6), designating the initial batch as the reference and incorporating the following covariates: disease status, sex, age, and hormone conditions. The effectiveness of the ComBat was validated using sample replicates between each batch in a matched pairs analysis. Analytes that exhibited significant differences post-correction were excluded from further analysis.

### Statistical analysis

We prospectively sought to enroll 50-100 participants to evaluate overall health and symptom changes. However, the study was terminated early due to an inability to reach the target sample size given that few people with Long COVID were vaccine naïve. Our final cohort comprised participants who met eligibility, completed the baseline survey, and were vaccinated at least once.

Cohort characteristics were reported as frequencies with proportions or medians with ranges. The overall health condition of participants after vaccination compared to before vaccination was described as the proportion of individuals with each response (i.e., better, worse, the same, don’t know) at each post-vaccination time point out of the number of individuals with a submitted survey at that time point.

For other symptom-related analyses, participants’ Likert scale responses to the physical and social effects associated with each symptom were coded numerically (**Supplementary table 1**). The proportion of symptoms experienced was calculated at each survey as the number of participants experiencing each symptom (i.e., symptom reported and non-zero response to the physical or social effect scales) out of the participants who completed the given survey.

The burden of each participant’s symptoms was summarized by summing across their responses to the symptom physical and social effect scales, separately, for each survey. Scores could range from 0 to 500 per survey (i.e., 125 symptoms per survey with a maximum score of 4). Higher values suggest greater symptom burden, and a value of 0 suggests no symptom burden. Changes in these values indicates a change in the number of symptoms experienced, the symptom severity, or both. We report the median, 2^nd^ and 3^rd^ quartiles, and range for each survey and effect (i.e., physical and social). Differences between surveys were not tested. Analyses were performed in R (v 4.2.2; R Foundation for Statistical Computing, Vienna, Austria).^35^

Differences in SARS-CoV-2 specific T-cell responses, anti-SARS-CoV-2 antibody responses measured by ELISA and REAP before and after vaccination were assessed using Wilcoxon matched-pairs signed rank tests. To assess correlation between observed T-cell responses and antibody levels as well as to determine concordance between the two different methods of determining anti-SARS-CoV-2 antibody levels, Spearman rank correlations were calculated. The correlation coefficients between assays were used to measure distances [1-absolute (correlation coefficients)], and hierarchical clustering was conducted using Morpheus.^36^. Participants were classified into outcome groups based on self-reported general health status before and after vaccination. The tests were all two-sided and Bonferroni-corrected P-values less than 0.05 were considered statistically significant.

Differences in SARS-CoV-2 T-cell responses, antibody levels, anti-viral antibody levels against common viruses and autoantibody levels among symptom outcome groups were also compared using Kruskal-Wallis tests. Further, to estimate the average differences in expression of each cytokine over the course of vaccination we used linear mixed models via Restricted Maximum Likelihood (REML) regression, estimating the cytokine expression over all three timepoints amongst three symptom outcome groups: those who did not improve or felt worse at weeks 6 and 12 post vaccination (n=3; Same/Worse), those who showed marginal improvement (n=2, Marginal [i.e. Better week 6; then Worse week 12]) and those who reported improvement (n=7, Better). The model incorporated a random effect for each individual as a random intercept, nested within their respective symptom outcome groups. The fixed effects in the model included the symptom outcome and time, along with an interaction term between them to investigate any potential modifying effect of time on the symptom outcome group. The analysis was conducted using the JMP statistical software platform (JMP^®^ Pro 17.0.0).

Statistical tests were performed using R (v 4.2.2)^35^, GraphPad PRISM(v 9.5.1), and JMP statistical software platform (JMP^®^ Pro 17.0.0).

### Machine Learning

Unsupervised hierarchical clustering was conducted on 162 plasma-derived analytes obtained from the multiplex proteomic assays to assess patterns of expression across the cohort using the JMP platform (JMP^®^ Pro 17.0.0). Data was standardized by factor and clustering was done based on Ward’s distance.

To further identify predictors of symptom improvement from the 162 plasma-derived analytes, we used Partial Least Squares (PLS) analysis via the Non-linear iterative partial least squares (NIPALS) algorithm with k-fold cross validation (k=5). The analysis was conducted using the JMP statistical software platform (JMP^®^ Pro 17.0.0). All plasma factors and sex were incorporated into the model. Final analysis involved reduction to 4 principal components, which simultaneously minimized the Van der Voet’s T-squared statistic (0.00, P=1.00) and the Root Mean PRESS (0.27) accounting for a sizeable portion of the variance in the data (cumulative pseudo-R-squared= 0.98). Post-analysis, the Variable Importance on Projection (VIP) score was generated for each feature and bootstrapped using Bayesian Bootstrapping. Bias-corrected 95% confidence intervals were calculated. Only features with 95% confidence intervals above the threshold cutoff of 0.8, corresponding to the standard threshold for importance,^37,38^ were considered significant.

## Results

Among 429 individuals screened between May 3, 2021 and February 2, 2022, 22 met inclusion criteria and consented to participate and 16 individuals completed the baseline survey and subsequently received a first dose of a COVID-19 vaccine; 14 completed all four surveys. People not enrolled had already received a vaccine, did not plan to be vaccinated, or were not able to travel to New Haven for biospecimen collection. The median age of the 16 included participants was 54 years (range 21-69), 13 (81%) were female, and 14 (88%) identified as Non-Hispanic White (**Table 1**). Immunophenotyping assays were completed on a subset of 11 out of 16. All participants reported that they tested positive for COVID-19 at least once with most reporting a PCR-based test (n=10, 62%).

**Table 1:** Baseline characteristics.

| Characteristic | n=16 (%) |
| --- | --- |
| Age, Median (Min-Max) | 54 (21-69) |
| Missing | 1 |
| Gender |  |
| Female | 13 (81%) |
| Male | 3 (19%) |
| Race/ethnicity |  |
| American Indian/Alaska Native | 1 (6%) |
| Hispanic | 1 (6%) |
| Non-Hispanic White | 14 (88%) |
| Tested for COVID-19 | 16 (100%) |
| Hospitalized due to COVID-19 | 3 (19%) |
| Hospitalized for COVID-19 or visited a hospital more than 2 weeks after infection | 4 (25%) |
| Test type |  |
| Antigen test | 2 (12%) |
| Not sure | 4 (25%) |
| PCR test | 10 (62%) |
| On your best days would you say you are ___ of health before COVID-19 |  |
| 0-25% of health before COVID-19 | 0 |
| 26-50% of health before COVID-19 | 1 (6%) |
| 51-75% of health before COVID-19 | 7 (44%) |
| 76-100% of health before COVID-19 | 8 (50%) |
| On your worst days would you say you are ___ of health before COVID-19 |  |
| 0-25% of health before COVID-19 | 5 (31%) |
| 26-50% of health before COVID-19 | 4 (25%) |
| 51-75% of health before COVID-19 | 6 (38%) |
| 76-100% of health before COVID-19 | 1 (6%) |

### Pre-vaccination health and symptoms

At baseline, on participants’ worst days, 9 (56%) felt they were 50% or less of their health before COVID-19. On participants’ best days, 7 (44%) reported feeling 51-75% of their health before COVID-19. The median number of symptoms per participant before vaccination was 23 (Q1-Q3, 13.8-27). The most frequently reported symptoms, in order, were brain fog (81%), fatigue (75%), difficulty concentrating (69%), difficulty sleeping (62%), heart palpitations (56%), shortness of breath or difficulty breathing (56%), anxiety (50%), memory problems (50%), dizziness (44%), feeling irritable (44%) (**Figure 1**). Three (19%) participants were previously hospitalized due to COVID-19 and 4 (25%) visited the hospital or were hospitalized for COVID-19 more than 2 weeks after onset of acute disease.

**Figure 1:**
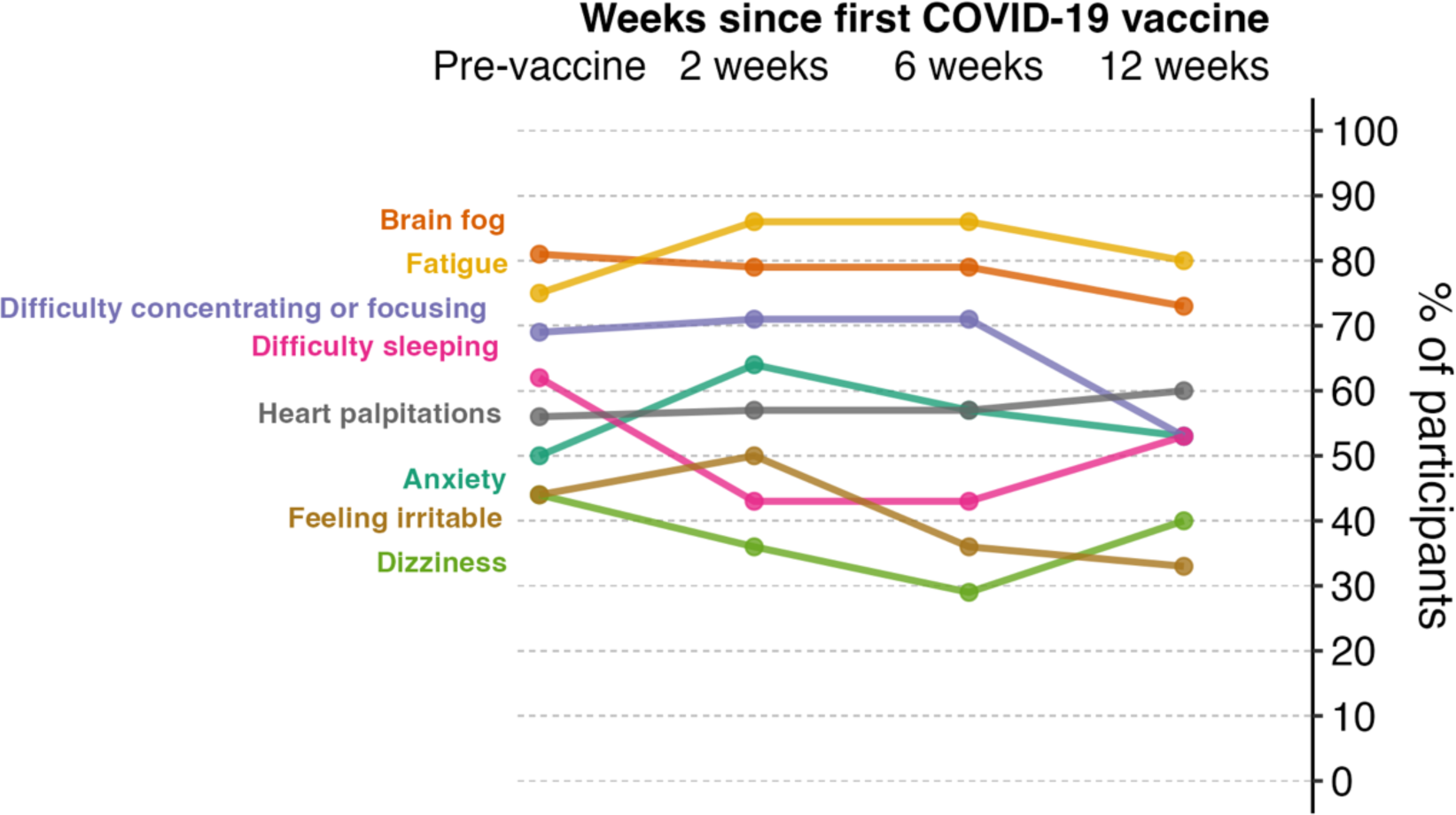
Trends of the ten most common symptoms before vaccination. The proportion of symptoms experienced was calculated at each survey as the number of participants experiencing each symptom (i.e., symptom reported and non-zero response to the physical or social effect scales) out of the participants who completed the given survey. Data missing for n=2 at 2 weeks and n=2 at 6 weeks.

### Post-vaccination changes in overall health

Eleven of 16 participants (69%) received the Pfizer-BioNTech vaccine (Comirnaty^®^), 3 (19%) received the Janssen vaccine as their first dose, and 2 (13%) received the Moderna vaccine (SpikeVax^®^). Nine of 13 participants (69%) recommended to receive a second dose in the primary series reported doing so (i.e., Janssen’s vaccine in the primary series was single dose). One participant was hospitalized for chest pain three days after receiving their first vaccine dose and again after their second dose.

Two weeks after vaccination, 6 out of 14 participants with completed surveys reported their health was better (43%), 3 (21%) said their health was the same, 1(7%) reported worse health, and 4 (29%) were not sure of a change (**Figure 2**). At 6 weeks after vaccination, 11 out of 14 (79%) said their health was better than before vaccination, 2 (14%) reported the same health, and 1 (7%) reported worse health. The participant with worse health 2 weeks after vaccination reported better health at 6 weeks. At 12 weeks, 10 out of 16 (62%) reported better health, while 3 (19%) reported the same health and 3 (19%) reported worse health. Two participants who reported better health at 6 weeks reported worse health at 12 weeks, which we classified as marginal improvement in subsequent analyses).

**Figure 2:**
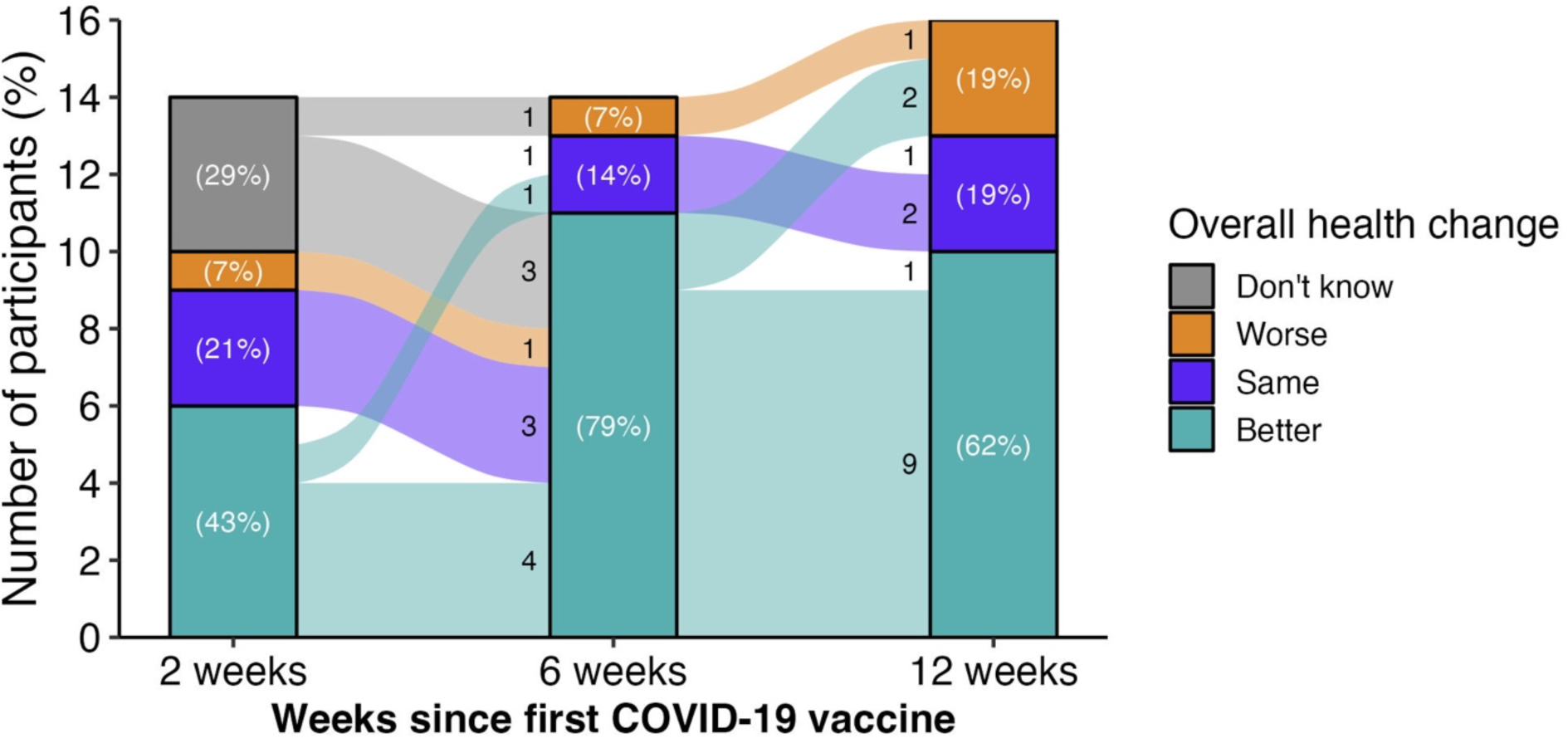
Overall health change since receiving first dose of COVID-19 vaccine, measured with surveys sent 2, 6, and 12 weeks after vaccination. Participants were asked “Would you say that your overall health, as compared to your health before the vaccine, is worse, better, or the same?” at each post-vaccination survey. Data missing for n=2 at 2 weeks and n=2 at 6 weeks.

The median number of symptoms per participant initially decreased from 23 (Q1-Q3 13.8-27, n=16) before vaccination to 19.5 (Q1-Q3 12-30.3, n=14) 2 weeks after vaccination, further declining to 17.5 (Q1-Q3 12.3-25.5, n=14) and 15.5 (Q1-Q3 12.5-24.5, n=16) at 6 and 12 weeks after vaccination, respectively.

Among the ten most common symptoms experienced at baseline, fatigue and brain fog remained common with 12 (75%) and 11 (69%) participants still reporting these symptoms 12 weeks after vaccination (**Figure 1**). Fewer participants reported difficulty concentrating or focusing at 12 weeks compared with before vaccination (8 [50%] at 12 weeks vs. 11 [69%] before vaccination). Other symptoms decreased modestly over time. The proportion of participants reporting fatigue (12 [75%]), heart palpitations (9 [56%]), and anxiety (8 [50%]) was the same at 12 weeks as before vaccination. The proportion of the ten most common symptoms at each survey are presented in **Supplementary table 2**.

Symptom burden appeared to decrease after vaccination on both physical and social effect scales (**Figure 3**). Before vaccination, the median physical effect score for all symptoms was 68.5 (Q1-Q3 37.5-84, range 13-138, n=16) and the median social effect score was 36.5 (Q1-Q3 14-51.5, range 1-84, n=16), where higher values represent worse symptom burden. Compared to before vaccination, the median physical effect score decreased to 46.5 (Q1-Q3 21.3-67.3, range 5-128, n=14) at 2 weeks after the first COVID-19 vaccine dose, then 36.5 (Q1-Q3 17.8-52.3, range 9-104, n=14) 6 weeks after vaccination, and 38.5 (Q1-Q3 15-49.5, range, 0-80, n=16) 12 weeks after vaccination. At 2 weeks after the first COVID-19 vaccine dose, the median social effect score decreased to 27.5 (Q1-Q3 7.3-34, range 1-80, n=14), then 23 (Q1-Q3 11-36.8, range 2-102, n=14) 6 weeks after vaccination, and 19 (Q1-Q3 3.8-27.5, range 0-61, n=16) 12 weeks after vaccination.

**Figure 3:**
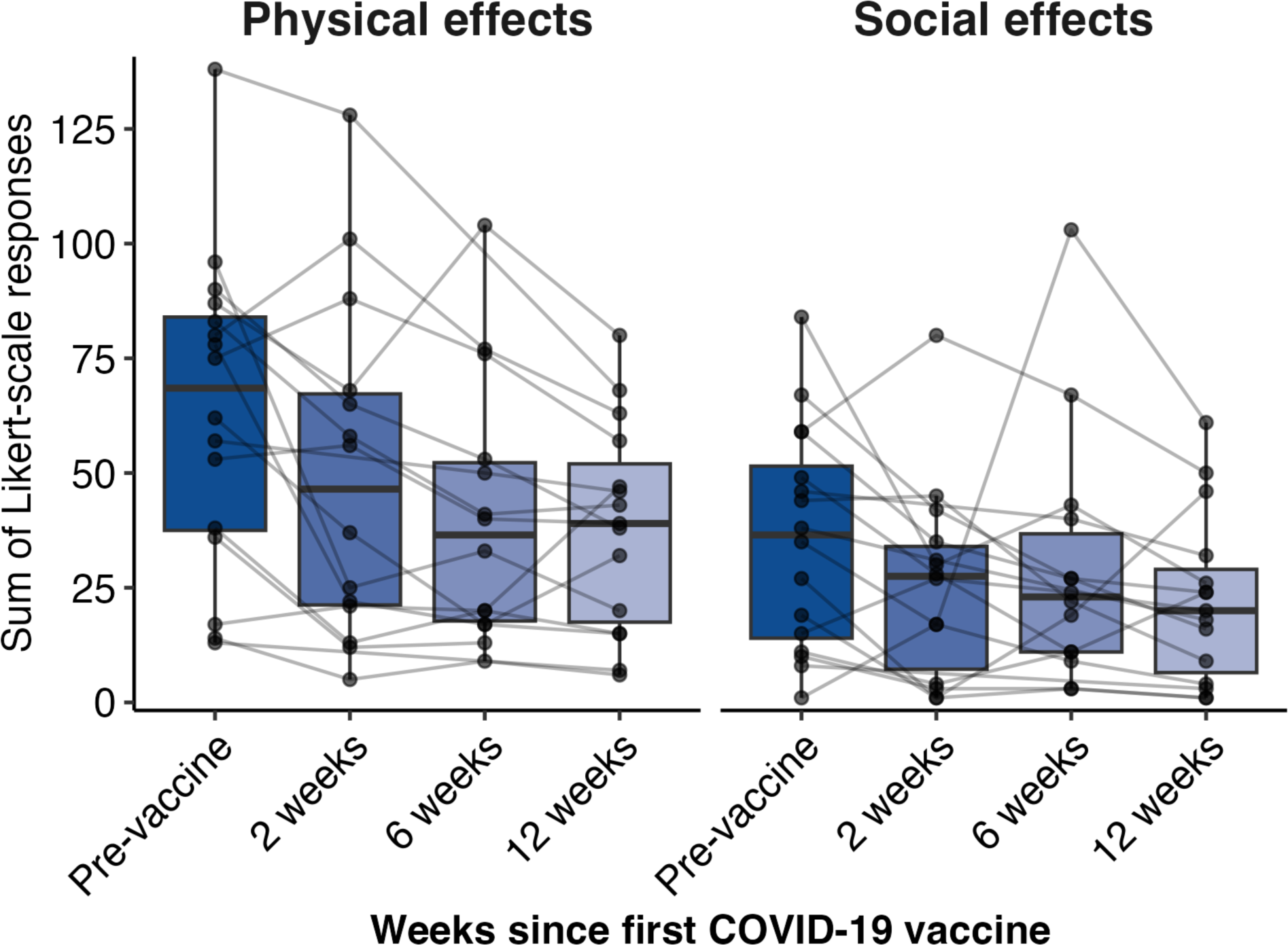
Distribution of the sum of participants’ responses to two measures of symptom severity—physical and social effects—measured before vaccination and surveys sent 2, 6, and 12 weeks after vaccination. To measure physical effect of each symptom from a list of 125 symptoms, participants were asked, “While experiencing these symptoms, how much do/did they bother you in terms of discomfort or pain?” Similarly, to measure social effects, participants were asked, “After quarantine, how much does/did the symptom impair your social or family functioning compared to pre-COVID? Responses for each symptom were scored 0 to 4 (Supplementary table 1) and summed for each participant. Boxplots show the distribution of responses, with points representing the score for each participant and lines showing participants’ trajectory across surveys. Data missing for n=2 at 2 weeks and n=2 at 6 weeks.

### SARS-CoV-2-specifc T-cells and antibody responses

To characterize the T-cell responses to SARS-CoV-2, sequencing of the CDR3 regions of T-cell receptor-β (TCR-β) chains was carried out. There was a significant increase in spike protein (**Figure 4a**; P_adjusted_=0.012, V1(**Figure 4b**; P_adjusted_=0.011) and V3 (**Figure 4c**; P_adjusted_=0.011) classifier scores at 6 weeks post-vaccination, which was indicative of an increase in SARS-CoV-2 specific T-cell clonal depth and breadth upon vaccination. By contrast and as expected, no significant differences were observed in classifier scores for non-spike protein TCRs with vaccination (**Figure 4d**; pre-vaccination vs 6 weeks: P_unadjusted_= 0.65; pre-vaccination vs 12 weeks: P_unadjusted_= >0.99). There were some individuals who retained high SARS-CoV-2 specific TCR clonality at 12 weeks post-vaccination, however the differences in model scores were not statistically significant in comparison with pre-vaccination. There was a significant decrease in V3 classifier score at 12 weeks post-vaccination as compared to 6 weeks (P_adjusted_=0.011), however this observation was not replicated using the V1 or spike specific classifier scores.

**Figure 4:**
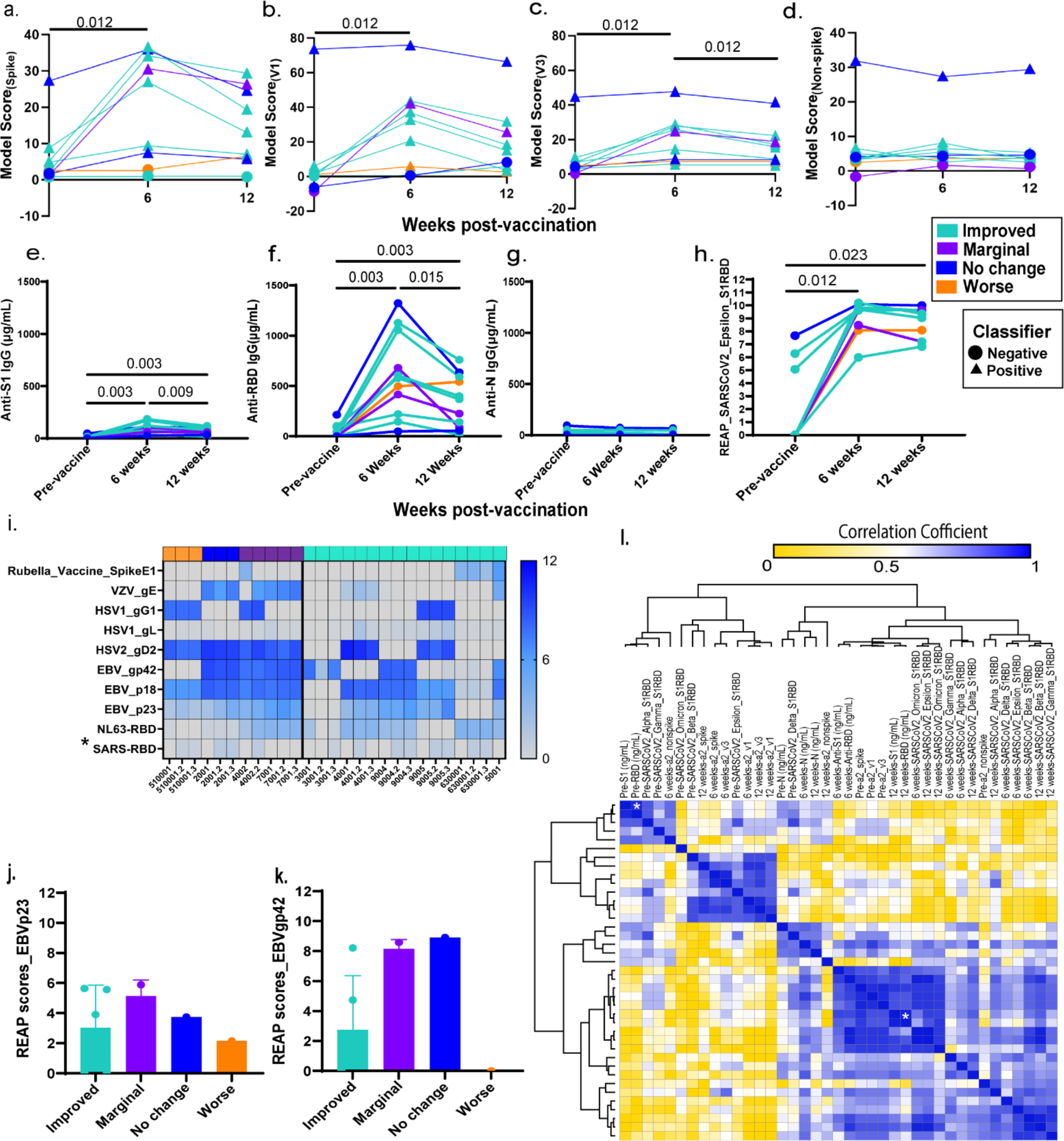
Vaccination resulted in increase in SARS-CoV-2 T-cell repertoires and specific humoral responses among Long COVID participants. (a) Model scores and binary classifications are plotted against days post-vaccination using spike viral protein specific classifier (b) COVID classifier version 1 (v1) (c) COVID classifier version 3 (v3) (d) Non-spike-specific protein classifier (e) Line plots of matched anti-SARS-CoV-2 S1 IgG concentrations before, 6 and 12 weeks post-vaccination in Long COVID participants. (f) Line plots of matched anti-SARS-CoV-2 RBD IgG concentrations before, 6 and 12 weeks post-vaccination in Long COVID participants. (g) Line plots of matched anti-SARS-CoV-2 N IgG concentrations before, 6 and 12 weeks post-vaccination in Long COVID participants. The color codes denote the reported health status at 6 and 12 weeks post-vaccination, better at both timepoints [teal], no change at both timepoints [blue], better at 6 weeks and worse at 12 weeks [purple] & worse at both timepoints [orange]. (h) Line plots of matched anti-SARS-CoV-2 Epsilon variant reactivity scores against the Spike protein assessed by Rapid Extracellular Antigen Profiling (REAP) (i) Heatmap of REAP reactivities against 10 viral proteins namely, proteins belonging to common viral pathogens from *Coronaviridae* (human SARS-CoV-1 viruses), *Herpesviridae* families, and the Rubella vaccine protein. Each protein and each participant timepoint are represented as a row and a column respectively. The participant IDs are mentioned below each column and the numbers after decimal denote the collection timepoints after vaccination (6 weeks= 2; 12 weeks= 3). Statistical significance determined by Wilcoxon Rank tests and corrected for multiple testing using the Bonferroni method. (j) EBV p23 REAP scores among outcome groups. Significance was assessed using Kruskal–Wallis tests. (k) EBV gp42 REAP scores among outcome groups. (l) Hierarchical clustering of Spearman Rank correlation coefficients of TCR model scores, antibody concentrations and REAP scores at all three timepoints. Only adjusted p-values of <0.05 are mentioned in line plots and denoted by asterisks in heatmaps.

Next, SARS-CoV-2 antibody responses were evaluated. A significant increase in anti-S1 IgG (**Figure 4e**; pre vs 6 weeks: P_adjusted_= 0.003, pre vs 12 weeks: P_adjusted_= 0.003) and anti-RBD IgG (**Figure 4f**; pre vs 6 weeks: P_adjusted_= 0.003, pre vs 12 weeks: P_adjusted_= 0.003) levels at 6 weeks and 12 weeks post-vaccination was observed without any observed significant rise in anti-N IgG levels (**Figure 4g**; pre vs 6 weeks: P_unadjusted_= 0.97, pre vs 12 weeks: P_unadjusted_= 0.37). The anti-S1 and anti-RBD IgG antibody levels peaked at 6 weeks (median anti-S1 IgG: 8.8×10^4^ ng/mL; median anti-RBD IgG: 5.0×10^5^ ng/mL) with a marginal decrease at 12 weeks (median anti-S1 IgG: 5.8×10^4^ ng/mL; median anti-RBD IgG: 2.2×10^5^ ng/mL). To further validate the humoral responses attributed to vaccination, SARS-CoV-2 spike protein reactivities were assessed using REAP. Participant antibody reactivities against Beta, Delta, Epsilon, and Omicron variant RBD epitopes were independently evaluated. A significant increase in reactivity across all non-Omicron RBD epitopes at 6 weeks post-vaccination (**Supplementary figures 1a-c**) and the Epsilon variant across 6- and 12-weeks post-vaccination (**Figure 4h**; pre vs 6 weeks: P_adjusted_= 0.011, pre vs 12 weeks: P_adjusted_=0.023) was observed.

### IgG responses to herpesviruses and autoantibodies to the extracellular proteome

Given that latent virus reactivation has been a hypothesis behind Long COVID pathobiology and evidence of recent Epstein-Barr Virus (EBV) reactivation has been reported,^30,39,40^ anti-viral REAP reactivities against two families of common viral pathogens namely, *Coronaviridae* (human SARS-CoV-1 viruses) and *Herpesviridae*, were assessed. Rubella vaccine spike antigen served as internal control as no changes were expected in reactivities with COVID-19 vaccination. As expected, there was a significant increase in REAP scores against SARS-COV-1 RBD upon vaccination at 6 weeks (**Figure 4i**; P_adjusted_= 0.05). This increase was maintained at 12 weeks, despite not being statistically significant after multiple testing correction (P_unadjusted_=0.031; P_adjusted_= 0.09). Herpesvirus reactivities varied across participants. However, no significant decrease in reactivities was observed post-vaccination among the herpesvirus antigens tested including EBV (**Figure 4i; Supplementary table 3**). Additionally, no differences in median reactivities were observed against EBV proteins p23 (P_unadjusted_=0.65) and gp42 (P_unadjusted_=0.06) across outcome groups at 6 and 12 weeks post-vaccination (**Figures 4j & 4k**).

Next, given prior reports of elevated autoantibodies targeting the exoproteome in severe acute COVID19,^41^ we assessed for changes in extracellularly targeted autoantibodies during vaccination (**Supplementary figure 2a**). No difference in the number of autoantibody reactivities at baseline (**Supplementary figure 2b**) or in the mean REAP score delta, representing the change in autoantibody magnitude over time (**Supplementary figure 2c**), between the groups was observed. Overall, autoantibodies were stable over time during vaccination (**Supplementary figures 2c-e**), with the mean REAP score delta close to 0 for all groups. These results are in concordance to a previous report focusing on autoantibody dynamics during SARS-CoV2 mRNA vaccination in healthy individuals without Long COVID.^42^

### Correlation between SARS-CoV-2 specific TCR and antibody levels

To further evaluate the relation between SARS-CoV-2 specific TCR scores with antibody levels and to assess the concordance among the orthogonal methods of antibody detection, correlation analyses were carried out. Three distinct clusters emerged when distances were calculated based on correlation values among TCR classifier scores and anti-SARS-CoV-2 antibody concentration as well as between ELISA and REAP assays at different timepoints. Each cluster indicated that there was a general concordance in antibody levels using orthogonal methods and TCR scores based on Spearman’s r (r_s_) and unadjusted p-values (**Figure 4l**, **Supplementary tables 4 and 5**). It was also observed that higher numbers of pre-vaccination SARS-CoV-2 specific TCR repertoire resulted in higher titers of antibodies both at pre-vaccination, 6- and 12-weeks post-vaccination along with an increase in spike protein specific TCR repertoire. Despite visually strong correlation patterns, due to the small sample size, only anti-SARS-CoV-2 S1 and anti-RBD antibody levels as detected by ELISA at pre-vaccination timepoint and at 12 weeks were statistically significant after multiple testing corrections (pre-vaccination: r_s =_0.96, P_adjusted_= 0.021; 12 weeks post-vaccination: r_s_ =0.98, P_adjusted_= 0.003; **Supplementary table 6**).

No significant differences were observed between post-vaccination increase in SARS-CoV-2 specific TCR classifier scores and improvement in overall health status [spike protein (P_unadjusted_=0.82), V1(P_unadjusted_=0.65), V3 (P_unadjusted_=0.56, and non-spike (P_unadjusted_=0.11)]. Similarly, no differences were also observed in self-reported health status and increase in anti-SARS-CoV-2 antibody levels [anti-S1 (P_unadjusted_=0.73), anti-RBD (P_unadjusted_=0.48) and anti-N (P_unadjusted_=0.94)].

### Soluble immune mediators

To understand the impact of vaccination on the cytokine, hormone, and proteomic profiles of individuals with Long COVID, unsupervised hierarchical clustering of 162 analytes measured in their plasma was first conducted (**Figure 5a**). Clustering analysis showed a consistent pattern in their plasma expression profiles at 6- and 12-weeks post-vaccination. Samples clustered by individual and not by timepoint post-vaccination, suggesting an entrenchment in the cytokine profile of each individual that was not significantly affected by vaccination.

**Figure 5:**
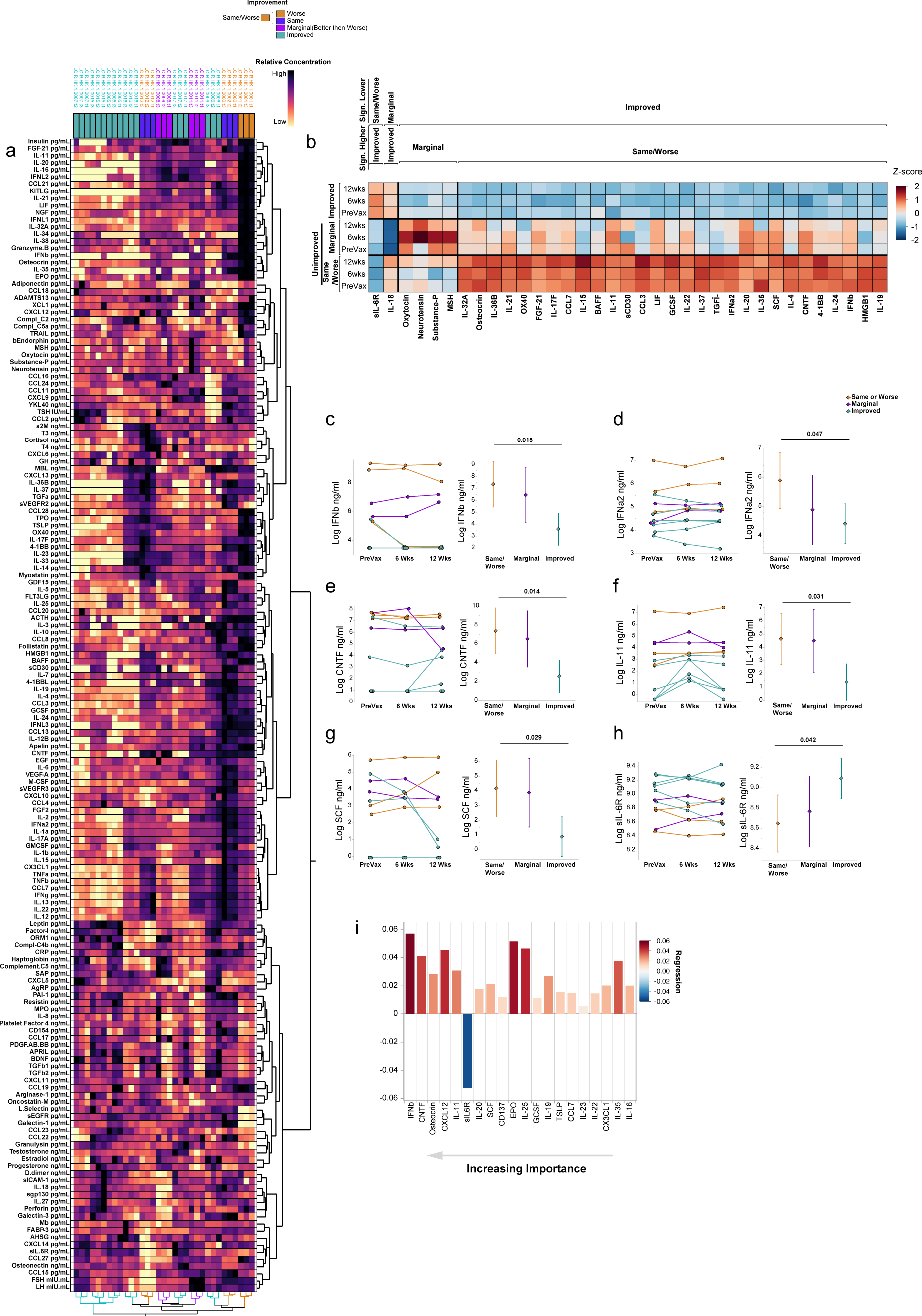
Elevated interferon and neuropeptide signaling is associated with poor recovery post-vaccination. (a) Unsupervised hierarchical clustering of plasma-derived analyte expression within the cohort for all three sample timepoints (pre-vaccination, 6 weeks post series completion, and 12 weeks post series completion). Color panel above heatmap shows the symptom outcome subgroup of each individual as indicated by the key. Samples for each individual are labeled by their sample code LC.R.HK.1.00XX.tX, where XX designates the patient ID and tX designates the timepoint (t1= pre vaccination, t2= 6 weeks post series completion, and t3= 12 weeks post series completion). Sample label color indicates further categorization into Same/Worse (orange), marginal improvement (i.e., better then worse; purple), and Improved (teal). Color scale is magma and is normalized for each analyte (data table columns) with darker colors indicating higher relative expression and lighter colors indicating lower expression as shown by the key. (b) Expression Heatmap of significant differentially expressed factors between symptom outcome groups (Same/Worse, Marginal, and Improved), as labeled. Each subgroup is further separated by the vaccine timepoint. Each factor was centered and standardized to generate a z-score and colors are representative of expression as indicated by the legend. To show significance between groups, samples were organized with outer brackets of the heatmap indicating the symptom outcome group demonstrating significantly lower expression and inner brackets indicating the comparator group from which significance is derived. Significance was determined using linear mixed models (LMM) via restricted maximum likelihood (REML) regression for log-transformed values, accounting for repeated measures across individuals over time as described in the methods and adjusted for multiple comparisons within each parameter using the Tukey method. (c-h) Example differentially expressed factors between symptom outcome groups as determined by LMM, previously described. (i) Top 20 bootstrapped predictors of symptom outcome (unimproved vs improved), determined by Partial Least Squares (PLS) optimized at eight components. Predictors are ordered by importance with highest importance on the left. Color and direction of each bar represents the relative regression association to unimproved individuals with positive values showing a positive association and negative values showing a negative association. Color is determined by regression as shown. Details of NIPALS and detailed results can be found in the methods and in extended data, respectively.

To understand the relationship of these plasma-derived analytes with post-vaccine symptom outcomes, the average expression levels of each analyte was compared over all three timepoints amongst three symptom outcome groups: those who did not improve or felt worse at weeks 6 and 12 post vaccination (n=3; Same/Worse), those who showed marginal improvement (n=2, Marginal [i.e. Better week 6; then Worse week 12]) and those who reported improvement (n=7, Better). To do so we constructed a linear mixed model using restricted maximum likelihood (REML) regression for each cytokine and accounted for both time and the interaction of time with each outcome group. Thirty-five factors were found to be significant amongst these subgroups, with the majority being significantly elevated in the Same/Worse group compared with the improved group. Most interferon factors we measured showed some elevation amongst the Same/Worse group, including IFN-β and IFN-α which were found to be significantly elevated in the Same/Worse group compared to the improved group (**Figures 5b-d**). Ciliary neurotrophic factor (CTNF; a neuropeptide that is released by the hypothalamus), IL-11, and SCF were also significantly elevated in the Same/Worse group compared to the improved group (**Figure 5e**). Other neuropeptides were noted to be elevated amongst the marginal group including oxytocin, neurotensin, substance P and MSH (**Figure 5b**). Notably, soluble IL-6 receptor (sIL-6R; an anti-inflammatory protein responsible for mitigating IL-6 signaling), was significantly higher amongst those who showed improvement compared to the Same/Worse group (**Figure 5h**).

We further employed Partial Least Squares (PLS) analysis with 5-fold cross validation on all 162 analytes to determine feature importance as predictors of symptom outcome and evaluate concordance with the significant features obtained from our LMM models. Final analysis involved reduction to 8 components, accounting for a sizeable portion of the variance in the data (cumulative pseudo-R-squared= 0.99). The top two significant predictors of the PLS analysis for non-improvement were IFN-β and CNTF respectively (**Figure 5i**). The top significant predictor of improvement was sIL-6R (**Figure 5i**), while sgp130, an important immunological partner to sIL-6R, was also associated with improvement, passing the initial VIP threshold criteria, though not the additional bootstrapping threshold criteria. Taken together these results suggested that high IFN and neuropeptide signaling were predictors of non-improvement while those involved in mitigating cytokine signaling, namely sIL-6R,was a predictor of improvement.

## Discussion

In this prospective cohort study of 16 vaccination-naïve individuals with Long COVID and significant symptoms at baseline, it was observed that most people improved or stayed the same during follow-up, but some experienced worsening. This study lacked concurrent controls and was small, so it is challenging to make definitive statements about the effect of vaccination, particularly since many people with Long COVID have fluctuations in their symptoms. However, the fact that symptom burden decreased on average and more people improved than worsened suggests that the vaccination was not overtly harmful. Future studies with controls are needed to understand the effect of vaccination on Long COVID symptoms.

Our findings are consistent with other studies and systematic reviews reporting improvement or non-significant change in self-reported health among people with Long COVID who were vaccinated for the first time.^18,43–45^ A single-center observational study in the United Kingdom identified 44 Long COVID patients (reporting a median of 4.1 and 3.6 symptoms per patient) who had received at least one dose of a COVID-19 vaccine and interviewed at 1 month and 8 months post-vaccination with the SF-36 and Warwick and Edinburgh Mental Wellbeing scores.^46^ After adjustment, health status measured with these instruments at 8 months did not differ compared to Long COVID patients who were not vaccinated. In an online cross-sectional survey study of 2,094 people in Switzerland, 35.5% of participants reported that their Long COVID symptoms improved, 28.7% reported their symptoms were stable, and 3.3% reported their symptoms worsened after vaccination.^47^ In a French target trial emulation study from the ComPaRe Long COVID cohort, COVID-19 vaccination was associated with a reduction in Long COVID severity and symptom burden at 120 days compared with those unvaccinated.^48^

Possible mechanisms of Long COVID have been proposed as: 1) a persistent viral reservoir or “viral ghost,” which are fragments of the virus (RNA, proteins) that linger after the infection has been cleared but are still capable of stimulating the immune system; 2) an autoimmune response induced by the infection; 3) reactivation of latent viruses; and 4) tissue dysfunction that results from inflammation triggered by the infection.^14,20^ Under these hypotheses, COVID vaccination may alleviate Long COVID symptoms through vaccine-induced T cells and antibody responses that may be able to eliminate the viral reservoir, and the “viral ghost,” diversion of autoreactive leukocytes, or removal of inflammatory sources leading to tissue dysfunction. Vaccination could also indirectly contribute to the control of latent virus reactivation by restoring proper T and B cell immunity against these herpesviruses.

Albeit with low sample sizes, this study provides evidence for alleviation of symptoms among Long COVID participants upon vaccination, along with an expected increase in SARS-CoV-2 specific T-cell repertoire and anti-SARS-CoV-2 spike protein specific IgG levels. However, clear results of hypothesis 1 (persistent viral reservoir) testing will be available once the results of the Paxlovid trials (NCT05595369^49^, NCT05668091^10^, NCT05823896^50^, NCT05576662^11^) and monoclonal anti-spike antibody (NCT05877508) are shared with the scientific community. In addition, a recent study did not find evidence of changes in circulating viral proteins in response to vaccination in those with Long COVID.^19^

Our study found that the plasma-derived soluble analyte profile showed a very stable pattern before and after vaccination, suggesting that vaccines had a minimal effect on the cytokine dynamics of individuals at least at the time points measured. Without concurrent controls, it is difficult to assess if this phenomenon is unique to individuals with Long COVID or is similar in controls. Nevertheless, an overall elevated cytokine pattern—namely in interferon and neuropeptide signaling— was detected among those who did not improve or showed only marginal symptomatic improvement post-vaccination. These findings pose an interesting observation that may help identify predictors of improvement versus non-improvement in larger studies. Elevated interferon signaling suggests the possibility of an ongoing infectious viral process in these individuals. The lack of improvement post-vaccination and the persistence of this signaling suggest that either the vaccine was incapable of producing the necessary antibodies and T cells that clear persistent infection when a viral reservoir exists, or that the main driver of disease in such individuals is not SARS-CoV-2, but re-emergence of a latent infection such as EBV or autoimmunity. More work will be needed to both confirm the findings of this small study and in turn to decipher a possible mechanism for elevation of interferon in these individuals, including the exploration of CNS involvement due to the elevation of neuropeptides which were also associated with poor improvement. A recent study has shown that the persistence of IFN signaling can lead to lower serotonin levels, a critical neurotransmitter^51^ which may also be involved in the symptom profile of individuals and or these outcomes. However, given the small sample size of our study, these possibilities can only be interpreted as speculative.

The limitations of this study include the lack of concurrent controls (i.e., individuals with Long COVID who remained unvaccinated or had a sham vaccination and completed the same surveys and provided biospecimens) to compare to our participants against and that most participants were recruited from an online Long COVID community and had to travel to New Haven, Connecticut for biospecimen collection and thus may not be representative of those with Long COVID. Participants had to be physically able to travel, so they may have been less likely to have severe Long COVID; at the same time, individuals more severely affected by Long COVID may have been more motivated to meet the travel requirements to participate in the study, as well as to be vaccinated. Participants also had to have the financial means and occupational flexibility to travel. Moreover, generalizability to all individuals with Long COVID cannot be determined especially for those who have developed Long COVID symptoms later in the pandemic (e.g., post-Omicron era). However, this study’s strengths are the prospective study design of vaccine naïve individuals with Long COVID, an increasingly rare population, with assessment of symptom burden, degree of physical and social disability, and immunophenotyping at multiple timepoints after vaccination.

In conclusion, in this study of 16 individuals living with Long COVID, most people improved or stayed the same, though some had worsening symptoms. Vaccination resulted in increase in SARS-CoV-2 specific T-cell populations and anti-spike protein IgG levels. The top predictor of participant non-improvement upon vaccination were IFN-β and CNTF, and sIL-6R was found to be a predictor of improvement. Future studies are needed to better understand the impact of vaccination in the health of people living with Long COVID.

## Supporting information

Participant survey

## Data Availability

All data produced in the present study are available upon reasonable request to the authors

## Acknowledgements

We want to thank the participants who contributed to the study and patient groups for their recruitment efforts and study design contributions. This work was supported in part by funds from Fred Cohen and Carolyn Klebanoff and by grants from National Institute of Allergy and Infectious Diseases (R01AI157488 to A.I.), the Howard Hughes Medical Institute Collaborative COVID-19 Initiative (to A.I.), and the Howard Hughes Medical Institute (to A.I.). J. J., T.T., and B. B. received research support from Yale University from the Food and Drug Administration for the Yale-Mayo Clinic Center of Excellence in Regulatory Science and Innovation (CERSI) (U01FD005938).

---

**Supplementary Figure 1:**
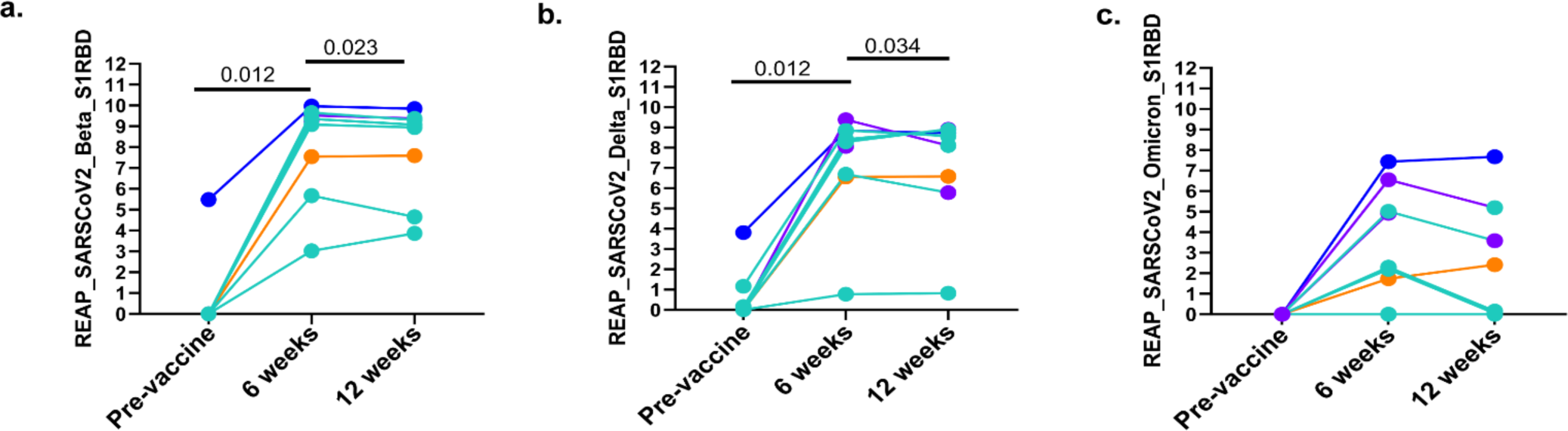
Increased reactivity in non-Omicron specific RBD epitopes post-vaccination. (a) Line plots of matched anti-SARS-CoV-2 Beta variant reactivity scores against the Spike protein assessed by Rapid Extracellular Antigen Profiling (REAP) (b) Anti-SARS-CoV-2 Delta variant reactivity scores (c) Anti-SARS-CoV-2 Omicron variant reactivity scores. Statistical significance was determined by Wilcoxon with Bonferroni correction for multiple comparisons. Only significant values are mentioned in the graphs.

**Supplementary Figure 2:**
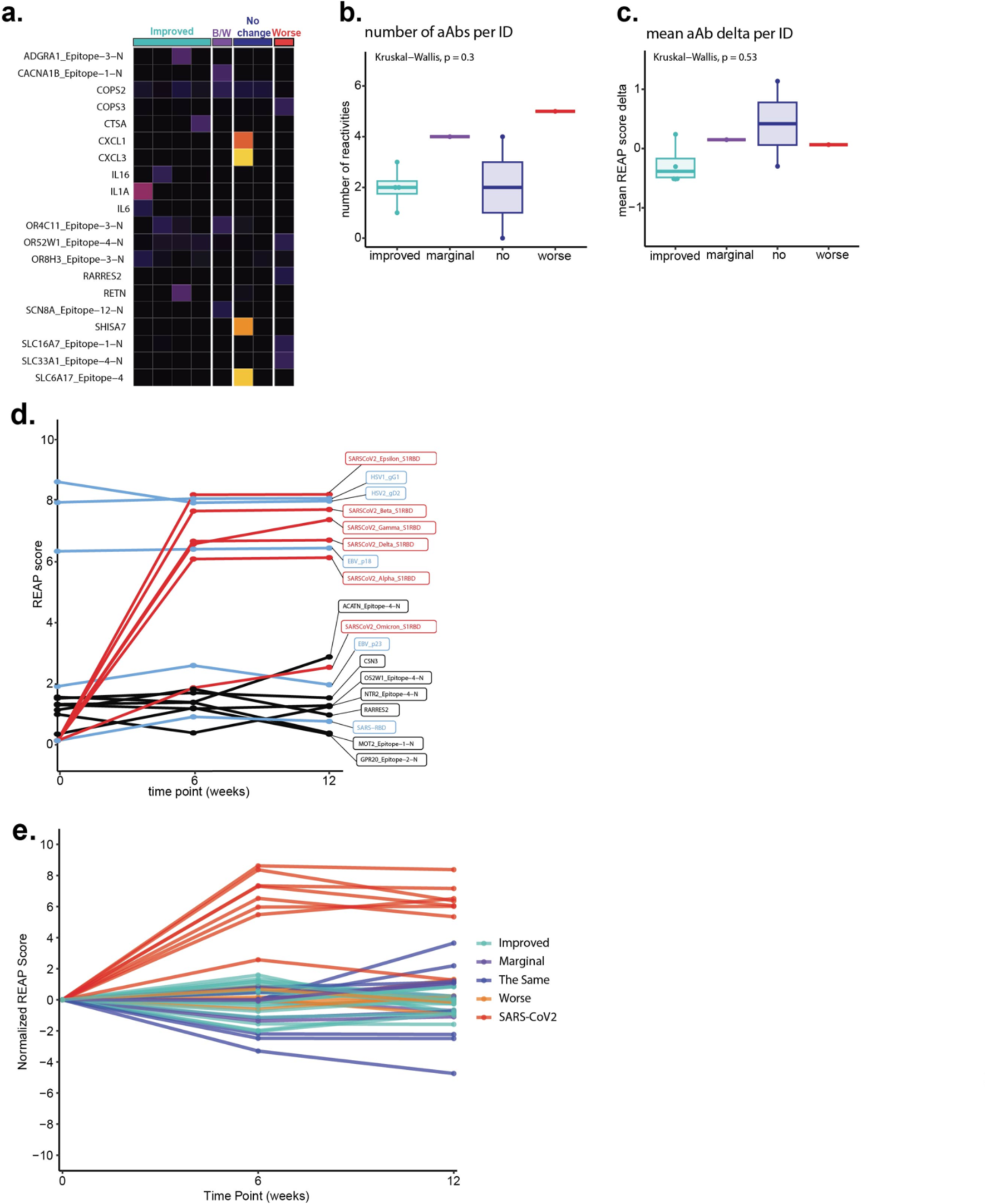
(a) REAP reactivities across the cohort at the first time point. Each column is one participant, grouped by cohort (B/W = marginal). Each row represents one protein. Reactivities shown have at least one participant with a REAP score ≥1. (b) The number of autoantibody (aAb) reactivities per individual (ID) by group. Significance was assessed using Kruskal–Wallis tests. (c) Average REAP score delta for autoantibody reactivities per individual from the first to the final time point. Significance was assessed using Kruskal–Wallis tests. For the box plots, the central lines indicate the group median values, the top and bottom lines indicate the 75th and 25th percentiles, respectively, the whiskers represent 1.5× the interquartile range. Each dot represents one individual. (d) Autoantibody trajectory for one individual during vaccination. Each line represents one reactivity (red = SARS-CoV2, blue = Herpesvirus, black = autoantibody. (e) overlay plot of all anti-viral and autoantibodies detected normalized to a starting REAP score of 0. Each line represents one reactivity.

**Supplementary Table 1:**
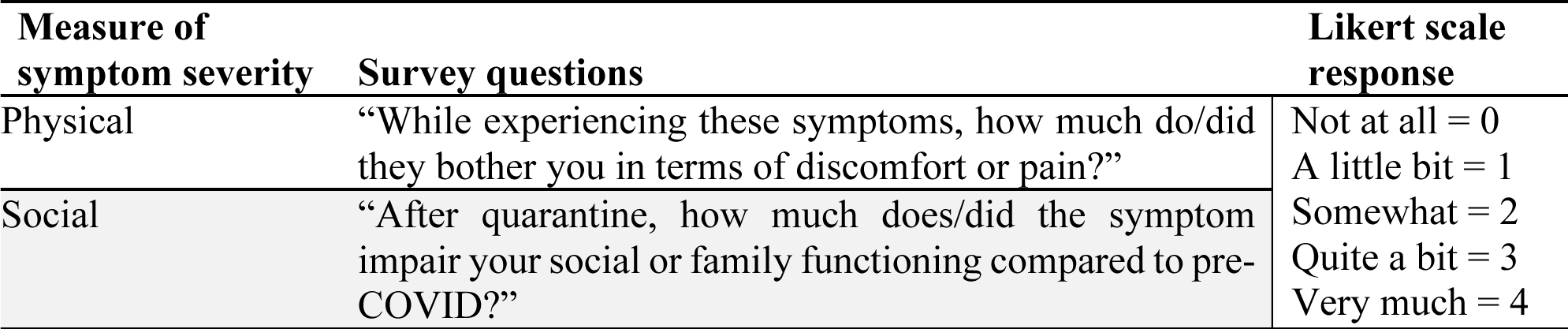
Survey questions assessing the physical and social effects for each symptom prior to vaccination and at surveys sent 2, 6, and 12 weeks after vaccination.

**Supplementary Table 2:** Ten most common symptoms prior to vaccination and at surveys sent 2, 6, and 12 weeks after vaccination. Data missing for n=2 at 2 weeks and n=2 at 6 weeks.

| Pre-vaccine (n=16) | 2 weeks (n=14) | 6 weeks (n=14) | 12 weeks (n=16) |
| --- | --- | --- | --- |
| Brain fog (81%) | Fatigue (86%) | Fatigue (86%) | Fatigue (75%) |
| Fatigue (75%) | Brain fog (79%) | Brain fog (79%) | Brain fog (69%) |
| Difficulty concentrating or focusing (69%) | Difficulty concentrating or focusing (71%) | Difficulty concentrating or focusing (71%) | Heart palpitations (56%) |
| Difficulty sleeping (62%) | Anxiety (64%) | Anxiety (57%) | Anxiety (50%) |
| Heart palpitations (56%) | Heart palpitations (57%) | Heart palpitations (57%) | Difficulty concentrating or focusing (50%) |
| Shortness of breath or difficulty breathing (56%) | Shortness of breath or difficulty breathing (57%) | Headache (50%) | Difficulty sleeping (50%) |
| Anxiety (50%) | Feeling irritable (50%) | Shortness of breath or difficulty breathing (50%) | Post-exertional malaise (50%) |
| Memory problems (50%) | Memory problems (50%) | Diarrhea (43%) | Headache (44%) |
| Dizziness (44%) | Post-exertional malaise (50%) | Difficulty sleeping (43%) | Inability to exercise or be active (44%) |
| Feeling irritable (44%) | Difficulty sleeping (43%) | Inability to exercise or be active (43%) | Shortness of breath or difficulty breathing (44%) |

**Supplementary File:** Blank study surveys sent to participants prior to vaccination and 2, 6, and 12 weeks after vaccination (PDF) and Supplementary tables 3-6 (Excel file).

